# Efficacy of cognitive behavioral therapy on non-motor symptoms and quality of life in Parkinson’s disease: A systematic review and meta-analysis

**DOI:** 10.1101/2020.06.12.20128967

**Authors:** Fangyi Luo, Mengfei Ye, Tingting Lv, Baiqi Hu, Jiaqi Chen, Junwei Yan, Anzhe Wang, Feng Chen, Ziyi He, Zhinan Ding, Jian Zhang, Hui Gao, Chao Qian, Zheng Liu

**Author notes:** Corresponding author at: Department of Behavioral Neurosciences, Medical Research Center, Medical College of Shaoxing University, Shaoxing, Zhejiang, China, 312000. These authors contributed equally to this work (Fangyi Luo, Mengfei Ye, Tingting Lv).

## Abstract

**Objective:** The aim of this study was to perform a quantitative analysis to evaluate the efficacy of cognitive behavioral therapy (CBT) on non-motor symptoms and its impact on quality of life (QOL) in Parkinson’s disease (PD).

**Methods:** We searched for randomized controlled trials in three electronic databases. Twelve studies, including 358 patients with PD, met the inclusion criteria. We determined the pooled efficacy by standard mean differences and 95% confidence intervals, using *I*^*2*^ to reveal heterogeneity.

**Results:** The result showed CBT had a significant effect on depression [-0.94 (95% CI, -1.25 to -0.64, *P* < 0.001)] and anxiety [-0.78 (95% CI, -1.05 to -0.50, *P* < 0.001)]. Moderate effect sizes were noted with stress [-0.60 (95% CI, -1.06 to -0.14, *P* = 0.01)] and sleep disorders [-0.44 (95% CI, -0.74 to -0.15, *P* = 0.003)]. There was no evident impact of CBT on fatigue or QOL. We found an intervention period > 8 weeks was advantageous compared with < 8 weeks, and CBT intervention was more effective than CBT developmental therapy.

**Conclusion:** We found that CBT in patients with PD was an efficacious therapy for some non-motor symptoms in PD, but not efficacious for fatigue and QOL. These results suggest that CBT results in significant improvement in PD and should be used as a conventional clinical intervention.

## Introduction

Parkinson’s disease (PD), a common neurodegenerative disorder with motor and non-motor symptoms that is second in prevalence to Alzheimer’s disease, affects more than 1% of the world’s population [1]. Compared with motor symptoms (resting tremor, bradykinesia, rigidity and gait disturbances), non-motor symptoms (mood disturbances, fatigue, apathy and sleep disorders) are often clinically underappreciated [2, 3]. Of these non-motor symptoms, the prevalence of mixed depression and anxiety reaches 50% and is more commonly found in patients with PD than in the general population [4]. Non-motor symptoms also have a negative effect on quality of life (QOL) for patients and their families, thus addressing these symptoms is urgent [5].

The treatment modalities for PD are expanding rapidly. Among all of the treatment options, pharmacotherapy remains first-line [6]. However, the side effects of long-term medication use are serious and in the presence of symptoms, frequent physical monitoring is needed which makes pharmacotherapy restrictive [7, 8]. Cognitive behavioral therapy (CBT) is a non-pharmacotherapy that has been shown to be effective for mental illness [9, 10]. Administration of regular sessions of CBT, which incorporates tailored interventions of relaxation training, thought monitoring and restructuring, sleep hygiene, worry control and others, has resulted in symptom improvements in patients with PD [11]. Koychev et al. [12] have reported the comprehensive benefits of CBT on the non-motor symptoms of PD in a clinical review, which assessed three psychiatric manifestations using four uncontrolled studies and two case series. Because of the barriers to receiving psychotherapy like CBT, such as lack of trained clinicians and fear of stigma, clinicians who have implemented the original form of CBT have encountered some obstacles [13, 14]. Therefore, CBT was further developed and more viable forms have been adopted widely that can be performed online rather than in person or may be administered in a group rather than with individuals. These changes may have led to inconsistencies in treatment outcomes.

Currently, several mate studies have respectively evaluated the efficacy of CBT in treating depression, anxiety and insomnia [15-17], but there has been no comprehensive quantitative analysis of the efficacy of CBT on non-motor symptoms of PD. The aim of this study was to perform a quantitative analysis of existing RCT trials to estimate the efficacy of CBT on non-motor symptoms and quality of life in PD, and ascertain which forms and duration of CBT intervention are best in an effort to inform clinical treatment.

## Methods

### Search methods

We performed a systematic search of RCTs on PubMed, Embase and the Cochrane Library up until January 2020. The following keywords and their synonyms were used: Parkinson’s disease, cognitive behavioral therapy, mindfulness-based cognitive therapy, acceptance and commitment therapy and dialectical behavior therapy. We attempted to access full texts of the retrieved literature.

### Inclusion and exclusion criteria

The inclusion criteria were as follows: (1) The study design was a randomized controlled trial (RCT), (2) participants had PD with any kind of non-motor symptoms, (3) the intervention was CBT and its derivative therapy, and (4) the outcome was evaluated by clinic recognition scales. The exclusion criteria were as follows: (1) case study, (2) lack of raw data or inadequate data and (3) no English publications or duplicated publications.

### Study screening and data extraction

Two authors (FL and TL) performed the search independently and excluded irrelevant articles according to the criteria above, and then accessed full-text articles where available. Both the authors evaluated the eligible articles, respectively, and gathered relevant information into a pre-designed data form. The form included author, year published, country of origin, study design, comorbidities, non-motor symptoms, mean age of patients, sample size and intervention for the two groups, measurement scales used, study duration and time to follow-up. Another data form was used for the outcome data gleaned from all of the scales used in the selected studies. We inverted the scales of the Parkinson’s Disease Questionnaire (PDQ), which measures QOL, to achieve consistency in the analysis so that high scores designated an improvement. Discrepancies were resolved by consensus discussion. When study details were incongruously documented between the two evaluating authors, a third author re-evaluated the study in question.

### Assessment of quality of literature

Two independent authors used the Cochrane Collaboration risk-of-bias assessment tool to detect the bias in the included papers, including random sequence generation, allocation concealment, blinding of participants and personal, blinding of outcome assessment, incomplete outcome data, selective reporting and other bias falling under the seven domains [18].

### Data analysis

The main analysis compared the differences between the two groups post-intervention to estimate the efficacy of CBT. Required data for analysis included means and standard deviations (SD). Data from subscales were combined according to the formula (Computational Formula). This study used the random-effects model. We obtained the pooled efficacy in standard mean differences (SMDs) and 95% confidence intervals using the various scales in the included studies, considering 0.2 as a small effect size, 0.5 as a moderate effect size and 0.8 as a large effect size. We reported comparability across the studies using *I*^*2*^ statistics to evaluate study heterogeneity. Subgroup and sensitivity analyses were performed to lessen the heterogeneity. We used State 12.0 for the graphs.

## Results

### Literature search

We found 1432 related articles through online databases, among which 369 repetitive articles were then excluded. We also excluded 968 nonconforming articles. Then, after reading through the titles and abstracts, we deleted 69 articles that did not meet the eligibility criteria. We acquired the full text of 26 articles and eliminated 14 of them for the specific reasons described in Fig. 1. Finally, 12 qualifying articles published between 2009 to 2019, comprising 358 participants, were analyzed in this meta-analysis.

**Fig. 1.**
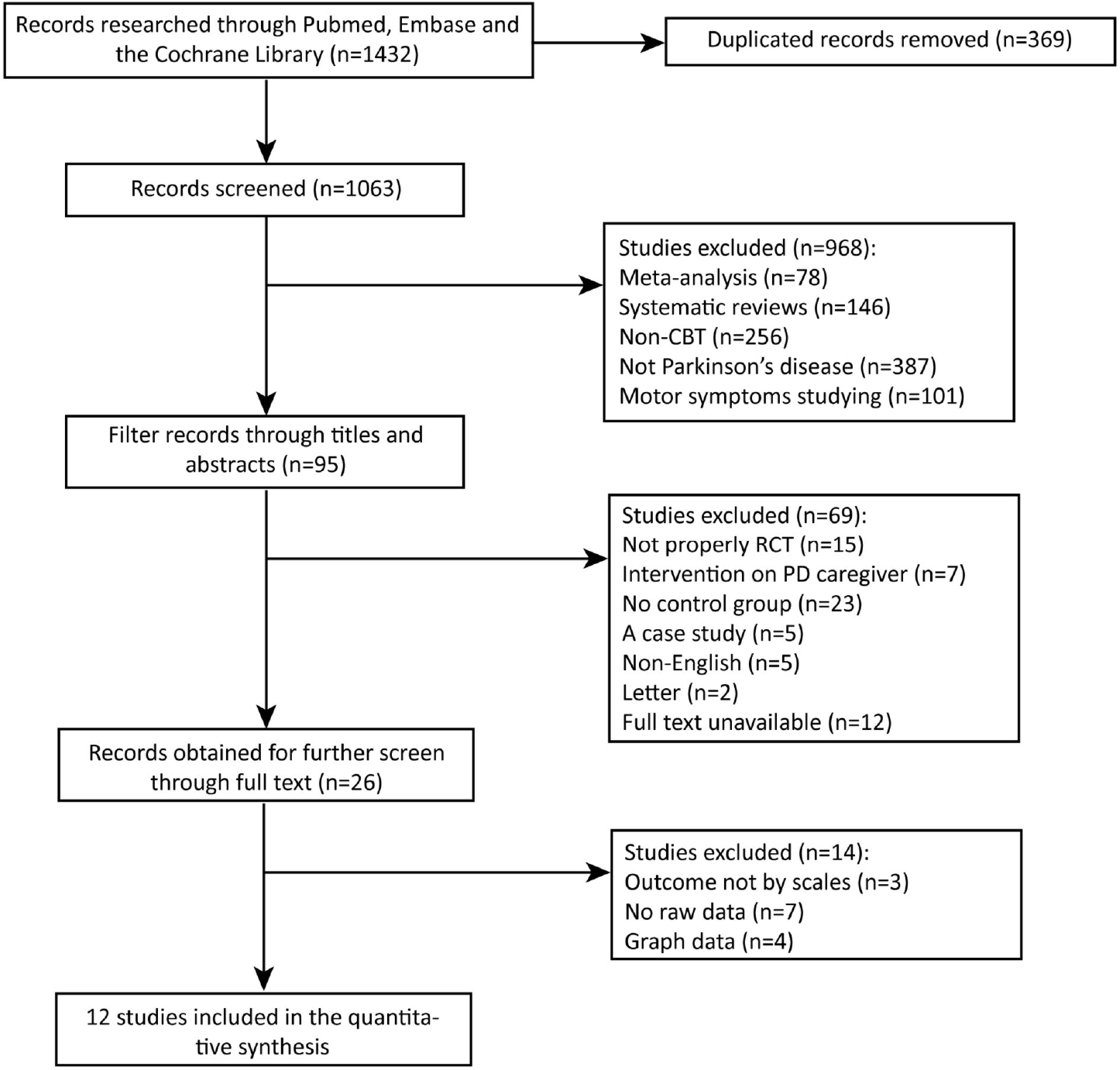
Screening process for trials included in the meta-analysis. Abbreviations: CBT = cognitive behavioral therapy; RCT = randomized controlled trial; PD = Parkinson’s disease.

### Characteristics of publications

The basic characteristics of the publications are shown in Table 1. Among the qualified articles, 10 studies performed CBT, one study performed Mindfulness-based Cognitive Therapy (MBCT) and one performed Acceptance and Commitment Therapy (ACT). The sample participants from Europe, Oceania and North America were 62.7 years old, on average, and comprised 33.4% women. Patients’ non-motor symptoms covered depression, anxiety, stress, sleep disorders (insomnia, sleep quality, and daytime sleep) and fatigue. The non-motor symptoms and outcome measure scales are displayed in Table 1. The biases of the included articles, based on the Cochrane Collaboration risk-of-bias assessment tool, are shown in Fig. 2.

**Table 1.** Main characteristics of studies.

| Author, Year | Country | Study Design | Comorbidity | Mean age | N (female%) | Intervention | Outcome | Duration |
| --- | --- | --- | --- | --- | --- | --- | --- | --- |
| Wuthrich et al. [39] (2019) | Australia | RCT (a single center study) | depression, anxiety | Total=68.8 | I=6; C=5 (36.4%) | I=telephone-delivered CBT<br>C=waitlist | GDS, GAI, WHO QoF | 10w |
| Berardelli et al. [40] (2018) | Italy | RCT | depression, anxiety, apathy | I=60.5<br>C=57.1 | I=9; C=9 (38.9%) | I=group CBT<br>C=psychoeducational | HAM-D, HAM-A, TAEC, PDQ8 | 12w |
| Patel et al. [41] (2017) | USA | RCT (a single center study) | insomnia, fatigue, daytime sleep, depression, | I=63.1<br>C=64.7 | I=14; C=14 (42.9%) | I=computerized CBT<br>C=standard sleep hygiene education | ISI, PIRS20, ESS, PHQ-9, FSS, PDQ8 | 6w |
| Hadinia et al. [36] (2017) | Switzerland | RCT | stress | I=65.0<br>C=67.0 | I=16; C=14 (23.3%) | I=group CBT<br>C=health enhancement program (HEP) | BELA, FKK, PDQ-39 | 9w |
| Calleo et al. [42] (2015) | USA | RCT | depression, anxiety | Total=62.9 | I=10; C=6 (12.5%) | I=CBT<br>C=enhanced usual care (EUC) | SIGH-D, SIGH-A | 12w |
| Troeung et al. [43]<br>(2014) | Australia | RCT | depression,<br>anxiety, stress | Total=66.0<br>I=68.0<br>C=62.0 | I=11; C=7<br>(33.3%) | I=CBT<br>C=waitlist | DASS-D, CCL-D,<br>DASS-A, CCL-A,<br>DASS-S, PDQ-39 | 8w |
| Romenets et al. [44]<br>(2013) | Canada | RCT (a<br>three-arm<br>study) | insomnia, sleep<br>quality, fatigue<br>depression, | I=64.5<br>C=69.5 | I=6; C=6;<br>(0.91%) | I=CBT/bright light therapy<br>C=placebo/bright light<br>therapy | SCOPA, ISI,<br>PDSS, PSQI,<br>BDI, KFSS,<br>PDQ-39 | 6w |
| Okai et al. [45]<br>(2013) | London | RCT | anxiety,<br>depression | I=59.3<br>C=57.9 | I=28; C=17<br>(31.1%) | I=CBT<br>C=waitlist | BDI, BAI | 6 months |
| Dobkin et al. [11]<br>(2011) | USA | RCT | depression, sleep<br>quality, anxiety,<br>negative thoughts | Total=64.6 | I=41; C=39<br>(40.0%) | I=CBT/clinical monitoring<br>C=clinical monitoring | HAM-D, BDI,<br>HAM-A, PSQI,<br>IQ, MSHS | 10w |
| Veazey et al. [46]<br>(2009) | USA | RCT | depression,<br>anxiety | Total=70.8 | I=5; C=5<br>(0%) | I=telephone-administered<br>CBT<br>C=support strategy | PHQ-9, BAI,<br>PDQ-39 | 8w |
| Rodgers et al. [47]<br>(2019) | Australia | RCT | depression,<br>anxiety | Total=63.7 | I=18; C=18<br>(44.4%) | I=MBCT<br>C=waitlist | GDS, DASS-D,<br>GA, DASS-A,<br>PDQ-39 | 8w |
| Ires et al. [48]<br>(2017) | Netherlands | RCT | depression,<br>anxiety | I=59.6<br>C=66.6 | I=19; C=19<br>(39.1%) | I=body awareness<br>intervention<br>C=treatment as usual<br>(TAU) | BDI, BAI,<br>PDQ-39 | 6w |

**Abbreviations:** BAI, Beck Anxiety Inventory; BDI, Beck Depression Inventory; BELA, the Burden Questionnaire for Patients with Parkinson's disease; CCL-A, Cognitions Checklist-Anxiety; CCL-D, Cognitions Checklist-Depression; DASS-A, Depression, Anxiety and Stress Scale-21-Anxiety; DASS-D, Depression, Anxiety and Stress Scale-21-Depression; DASS-S, Depression, Anxiety and Stress Scale-21-Stress; ESS, Epworth Sleepiness Scale; FKK, The Questionnaire for Disease-Related Communication; FSS, Fatigue Severity Scale; KFSS, Krupp Fatigue Severity Scale; GAI, Geriatric Anxiety Inventory; GDS, Geriatric Depression Scale; HAM-A, Hamilton Rating Scale for Anxiety; HAM-D, Hamilton Rating Scale for Depression; IQS, Inference Questionnaire Score; ISI, Insomnia Severity Index; PDQ, The Parkinson's Disease Questionnaire; PDSS, Parkinson's Disease Sleep Scale; PHQ-9, Patient Health Questionnaire; PIRS20, Pittsburgh Insomnia Rating Scale; PSQI, Pittsburgh Sleep Quality Index; SCOPA, Scales for Outcomes in Parkinson's Disease-Night; SIGH-A, Structured Interview Guide for the Hamilton Anxiety Scale; SIGH-D, Structured Interview Guide for the Hamilton Depression Scale; TAEC, The Apathy Evaluation Scale; WHO QoF, World Health Organization Quality of Life-Brief.

**Fig. 2.**
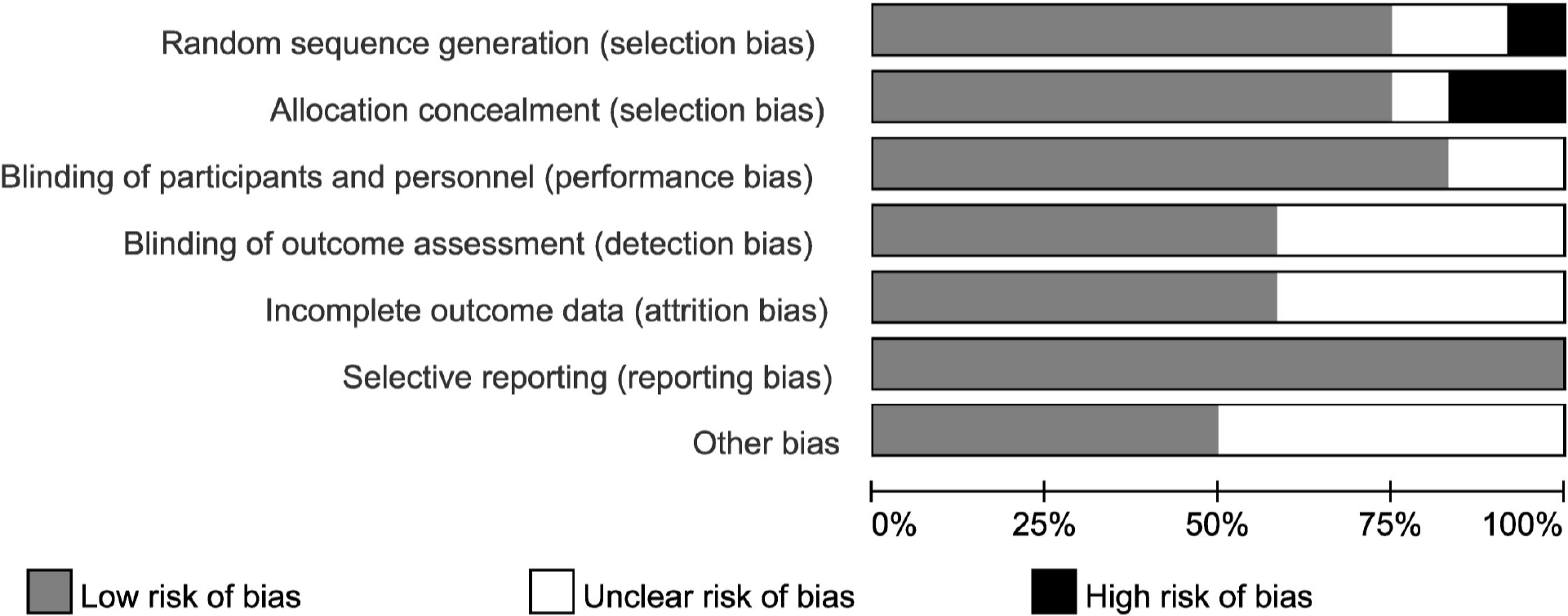
Risk of bias in the included trials.

### Synthesis of results

We evaluated the non-motor symptoms with the provided data at post-treatment. Patients in the eleven studies reporting depression displayed large improvement in depression (−0.94, 95%CI -1.25 to -0.64, *P* < 0.001, *I*^*2*^ = 42%) after intervention (Fig. 3A). Significant effects of CBT were also found in anxiety (−0.78, 95%CI -1.05 to -0.50, *P* < 0.001, *I*^*2*^ = 0%; Fig. 3B) and stress (−0.60, 95%CI -1.06 to -0.14, *P* = 0.01, *I*^*2*^ = 0%; Fig. S1) as reported by eight and two studies, respectively. There were three studies related to sleep disorders in which CBT was noted to have had a moderate effect (−0.44, 95%CI -0.74 to -0.15, *P* = 0.003, *I*^*2*^ = 0%) upon completion of the study (Fig. 4). There was no evident impact of CBT on fatigue (−0.22, 95%CI -0.92 to 0.49, *P* = 0.55, *I*^*2*^ = 0%; Fig. S2) and quality of life (0.08, 95%CI -0.24 to 0.40, *P* = 0.63, *I*^*2*^= 30%; Fig. 5). There was a set of data about apathy (−2.50 vs. 1.60), negative thoughts (−1.95 vs. -0.73) and psychiatry rating (−6.20 vs. 0.3) showing that the intervention group had more improvement than the control group (Figs. S3, 4 and 5). Within all of the symptoms the heterogeneity was 0%, except for depression and QOL, with a heterogeneity of 42% and 30%, respectively. One study was removed from the assessment of anxiety because it increased the *I*^*2*^ value to 69%. Our rationale for this exclusion is described below.

**Fig. 3.**
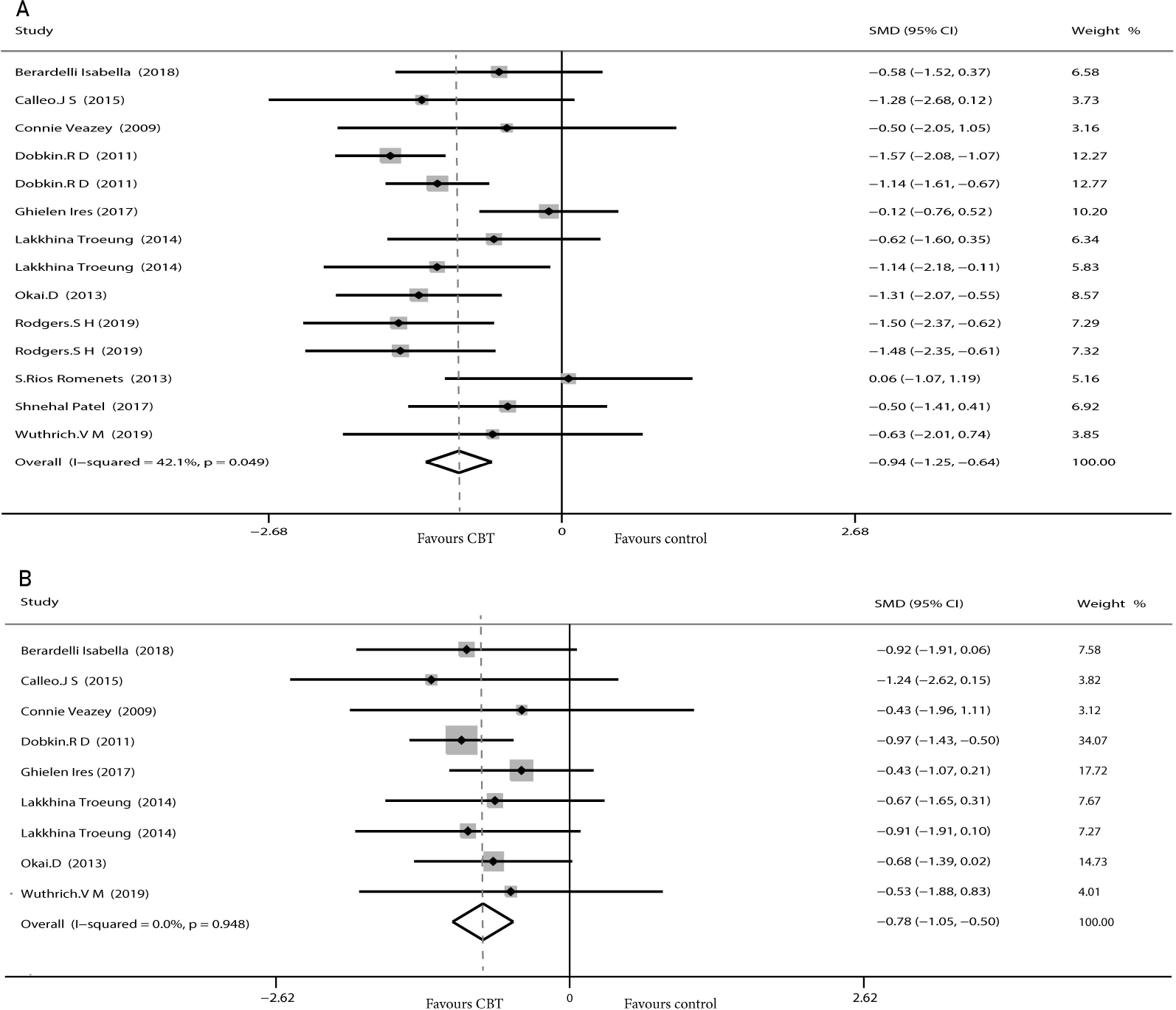
Forest plot of random effects model meta-analysis of the effect of CBT on depression and anxiety. Abbreviations: CBT = cognitive behavioral therapy; A = synthesis effect of depression; B = synthesis effect of anxiety.

**Fig. 4.**
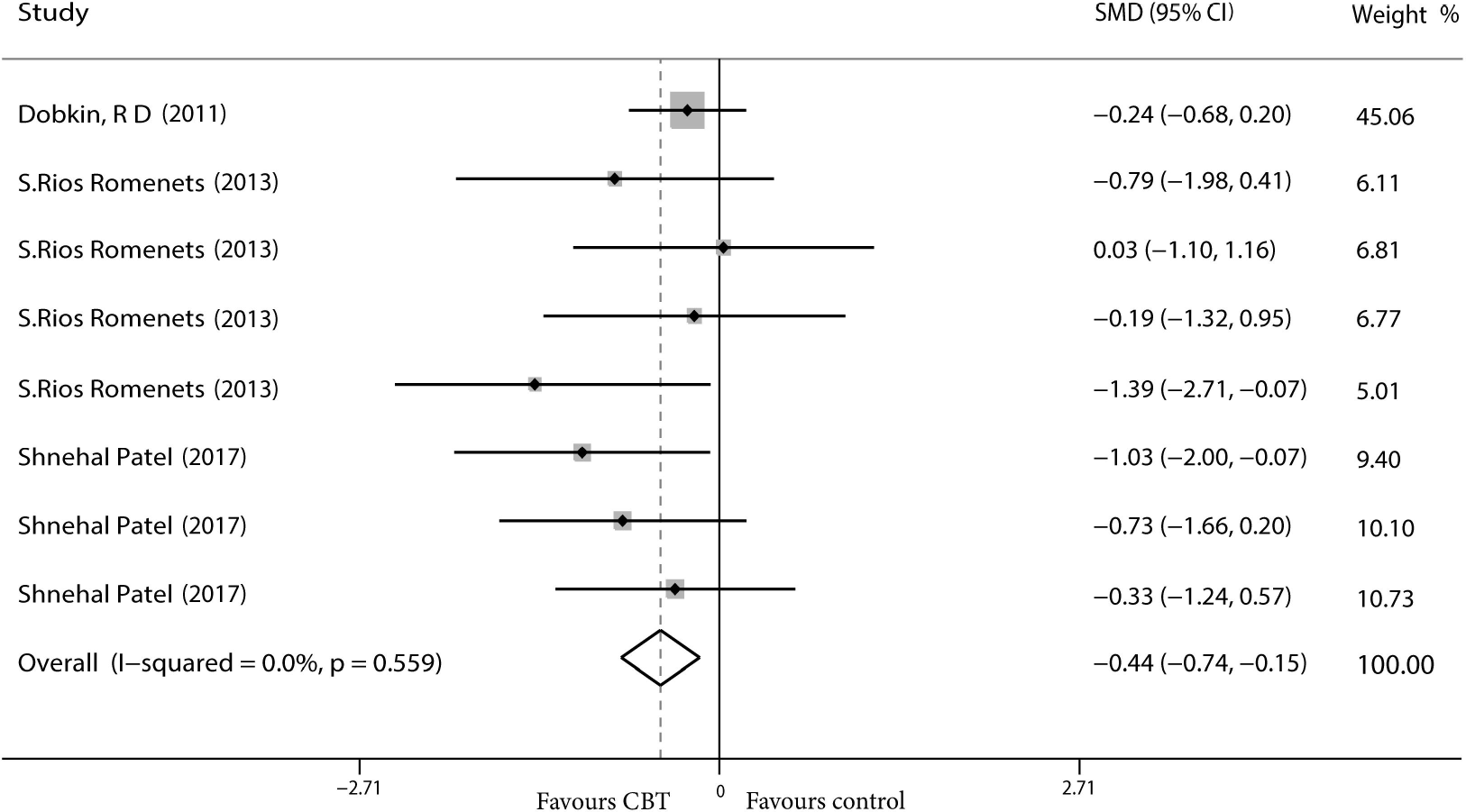
Forest plot of random effects model meta-analysis of the effect of CBT on sleep disorders. Abbreviations: CBT = cognitive behavioral therapy.

**Fig. 5.**
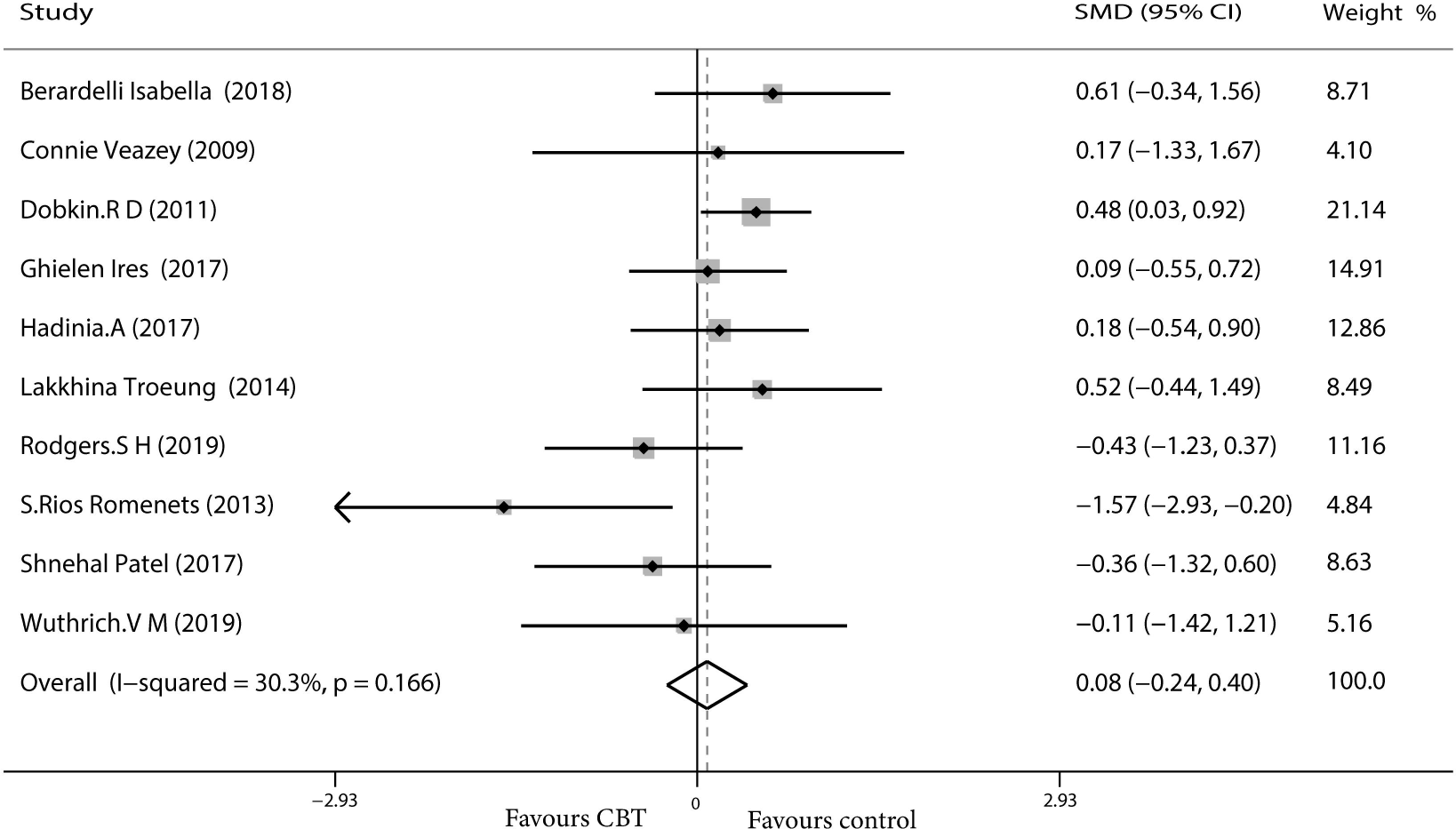
Forest plot of random effects model meta-analysis of the effect of CBT on QOL. Abbreviations: CBT = cognitive behavioral therapy; QOL = quality of life.

### Subgroup analysis

We performed a subgroup analysis of depression, anxiety, sleep and quality of life, based on the nationality of participants, duration of intervention and type of CBT intervention administered. When analyzing online (telephone or computer) and offline (in-person) interventions, we excluded two studies that used both methods. Stress and fatigue were not included as symptoms in the final analysis because the relevant studies lacked sufficient data.

#### Depression

Duration of CBT intervention was divided into < 8 weeks (−0.19, 95%CI -0.66 to 0.28, *P* = 0.43, *I*^*2*^ = 0%), 8 to 10 weeks (−1.16, 95%CI -1.6 to -0.72, *P* < 0.001, *I*^*2*^ = 0%) and ≥ 10 weeks (−1.24, 95%CI -1.52 to -0.95, *P* < 0.001, *I* ^*2*^= 0%; Fig. 6A). Our results showed that more than 8 weeks of intervention was more advantageous. The effects of all three intervention durations were similarly large (Fig. 6B). Intervention online (−0.53, 95%CI -1.21 to 0.51, *P* = 0.13, *I*^*2*^ = 0%) had a lower effect than offline (−0.84, 95%CI -1.28 to -0.40, *P* = 0.0002, *I*^*2*^ = 50%), but the latter had moderate heterogeneity (Fig. 6C). Individual intervention (−1.25, 95%CI -1.50 to -0.99), *P* < 0.001, *I*^*2*^ = 0%) resulted in greater improvement, compared with group intervention (−0.41, 95%CI -0.81 to -0.01, *P* = 0.04, *I*^*2*^ = 0%; Fig. 6D).

**Fig. 6.**
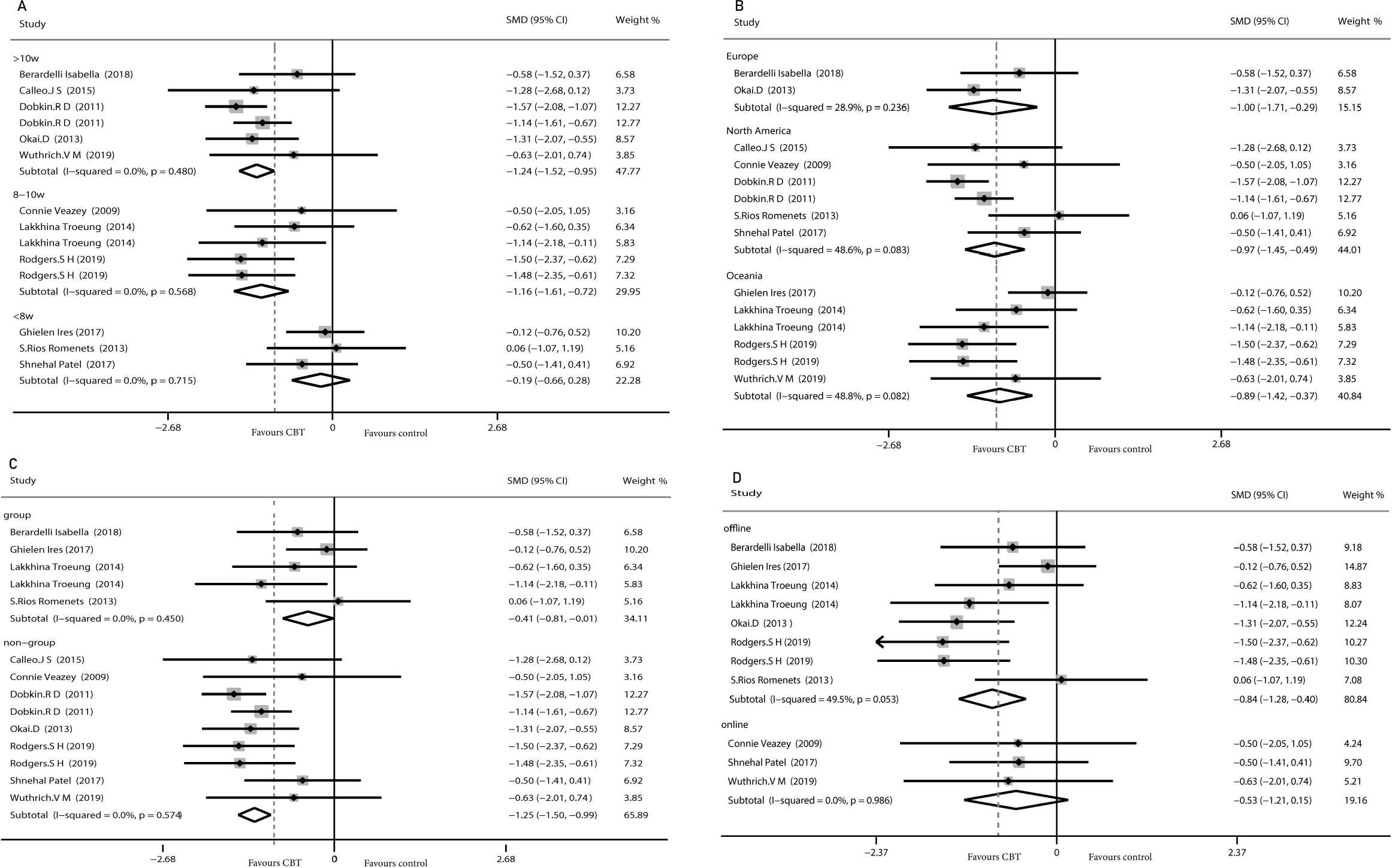
Forest plot of random effects model meta-analysis of the effect of CBT on subgroups of depression. Abbreviations: CBT = cognitive behavioral therapy; A = the effect of intervention time; B = the effect of participants’ regions; C = the effect of group or non-group intervention; D = the effect of online or offline intervention.

#### Anxiety

Some adjustments were made to the time division of the intervention to ensure the data were analyzable. The result showed intervening for 8 to 10 weeks (−0.92, 95%CI-1.36 to -0.48, *P* < 0.001, *I* ^*2*^ = 0%) obtained the optimum reduction in anxiety (Fig. 7A). North American patients had the greatest improvement in anxiety (−0.95, 95%CI-1.37 to -0.53, *P* < 0.001, *I*^*2*^ = 0%) compared with those from Europe (−0.76, 95%CI-1.34 to -0.19, *P* = 0.009, *I*^*2*^ = 0%) and Oceania (−0.59, 95%CI -1.03 to -0.14, *P* = 0.01, *I*^*2*^ = 0%; Fig. 7B). Intervention offline (−0.66, 95%CI -1.03 to -0.30, *P* = 0.0004, *I*^*2*^ = 0%; Fig. 7C) and in person (−0.86, 95%CI -1.21 to -0.51, *P* < 0.001, *I*^*2*^ = 0%; Fig. 7D) were more useful than the other method.

**Fig. 7.**
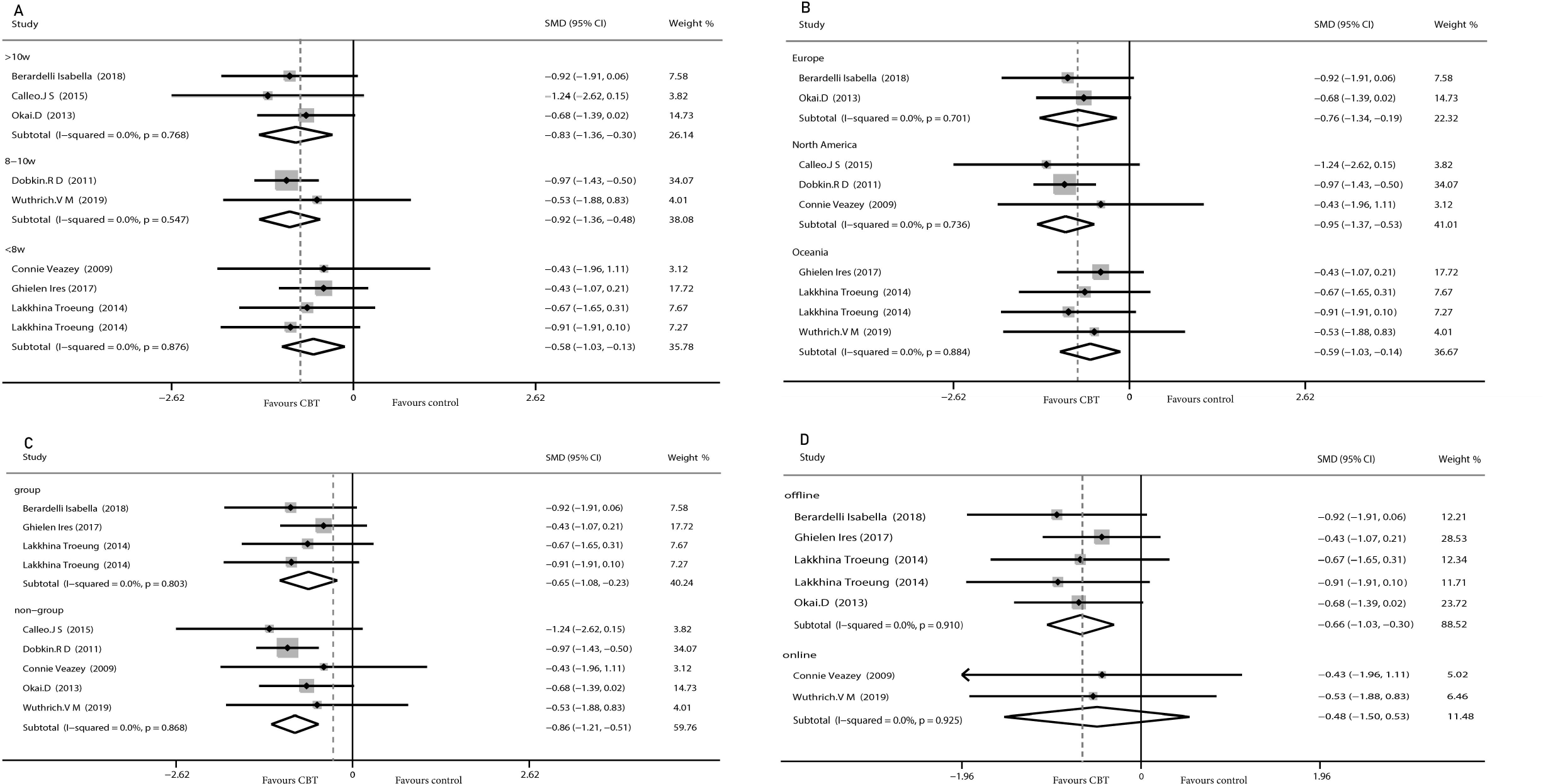
Forest plot of random effects model meta-analysis of the effect of CBT on subgroups of anxiety. Abbreviations: CBT = cognitive behavioral therapy; A = effect of intervention time; B = effect of participants’ regions; C = effect of group or non-group intervention; D = effect of online or offline intervention.

#### Sleep

Patients who participated in sleep assessments were all from North America and were only treated for 6 or 10 weeks. Online CBT offered preferable efficacy (−0.68, 95%CI-1.22 to -0.15, *P* = 0.01, *I*^*2*^ = 0%; Fig. 8A), and no significant difference between individual and group intervention was found (Fig. 8B), with both showing medium effect. Sleep covered two items, insomnia (−0.94, 95%CI -1.48 to -0.41, *P* = 0.0005, *I*^*2*^= 0%; Fig. 8C) and sleep quality (−0.20, 95%CI -0.59 to 0.18, *P* = 0.30, *I*^*2*^ = 0%; Fig. 8D). We found that CBT had evident improvement on insomnia with no increase in sleep quality.

**Fig. 8.**
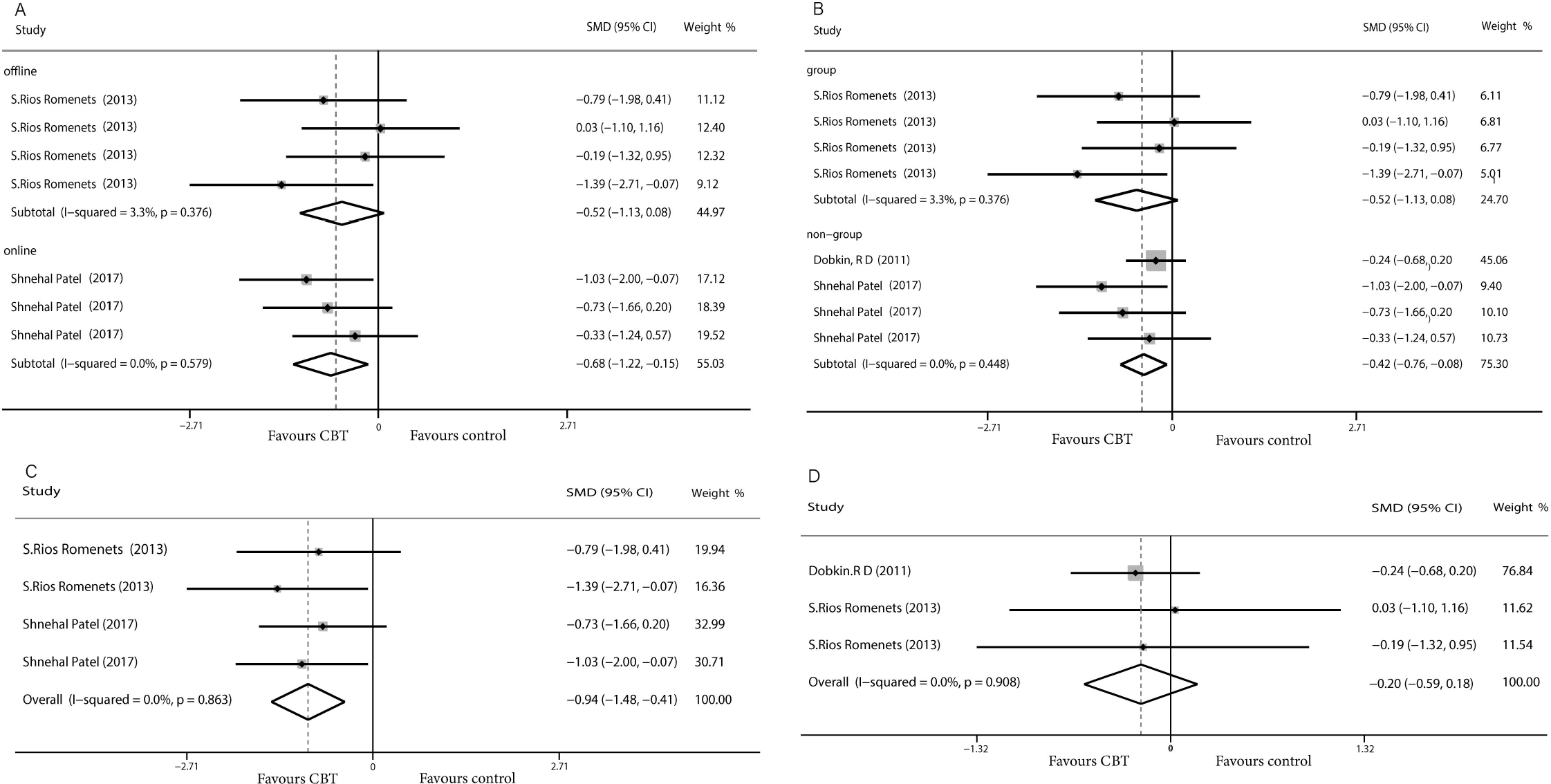
Forest plot of random effects model meta-analysis of the effect of CBT on subgroups of sleep disorders. Abbreviations: CBT = cognitive behavioral therapy; A = effect of group or non-group intervention; B = effect of online or offline intervention; C = insomnia; D = sleep quality.

#### Quality of Life

The pooled results on the effect of CBT on QOL in patients with PD had 30% heterogeneity, and we tried to discover the source of heterogeneity and mitigate its effect. We obtained a more reliable result after removing one article (0.21, 95%CI-0.05 to 0.46, *P* = 0.12, *I* ^*2*^= 0%), although the result remained insignificant. Only interventions lasting more than 10 weeks were considered to have a moderate effect on QOL (0.45, 95%CI 0.06 to 0.83, *P* = 0.02, *I*^*2*^ = 0%; Fig. S6A). No significant results were found by regions and by intervention methods. We analyzed the scales used and found moderate effects were reported using scales other than the PDQ (0.42, 95%CI -0.01 to 0.84, *P* = 0.05, *I*^*2*^ = 0%), while we found no efficacy of CBT in reducing QOL when measured with the PDQ (Fig. S6B).

## Discussion

Existing data have shown that a variety of non-motor symptoms appear in PD before motor features and even before diagnosis by several years [19, 20]. Thus, the symptoms of PD are present throughout the disease course and can have an impact on each of its stages. Symptoms in patients with PD who have higher rates of co-morbid psychiatric and executive dysfunction usually differ from symptoms in patients with non-PD psychiatric disorders [21]. Thus, the use of original CBT in patients with PD may have been inappropriate, so CBT protocols have been modified to treat psychological problems in this population. In this study, we upgraded prior systematic reviews and meta-analyses by including existing eligible RCT studies, demonstrating that CBT had a considerable efficacy on non-motor symptoms of PD.

There is a large body of literature on depression and anxiety due to their high incidence rates in Parkinson’s disease [22, 23]. According to the data we collected, CBT greatly reduced the scores for depression (*E* = -0.94) and anxiety (*E* = -0.78). One article by Rodgers et al. with a high level of heterogeneity was excluded from the analysis of anxiety because the authors used two scales (DASS-21 and GAI) to assess changes in anxiety, with inconsistent results. The authors suggested that these inconsistencies may have been related to the low baseline level of anxiety in their patients and the type of intervention implemented. We also performed a subgroup analysis on the duration of CBT intervention, which revealed that > 8 weeks of CBT was more effective than < 8 weeks. We can assume the longer the intervention, the better the outcome. PD is a persistent disease with chronic symptoms, making rapid treatment response difficult, especially because most patients with PD are older [24]. CBT programs generally lasted between 6–12 weeks in our included studies, excepting one that was 6 months long. Therefore, long-term intervention RCT trials are needed to further explore the effects of CBT. Bomasang-layno et al. [25] have reported consistent results, but they only included five relevant RCTs with limited data. In contrast, our results would be more stable and reliable.

When analyses were narrowed to the trials in offline interventions, effect sizes appeared to be large compared with no efficacy found with online (telephone or computer) intervention analyses. Two studies (by Calleo and Dobkin) that included both online and offline CBT approaches were excluded from the subgroup analysis. Eligible studies of online CBT had small sample sizes and one study reported a high drop-out rate, which may have altered our results. Problems with face-to-face CBT include limitations in clinician-patient interaction due to patients’ physical disability and the time patients spent traveling to appointments [26]. These barriers may have contributed to the development of telemedicine. The accessibility of telemedicine reduces the burden and psychological distress of face-to-face CBT and appears to lower attrition [27, 28]. Telemedicine is a promising option for the treatment of non-motor symptoms in PD.

We also limited our focus on the trials with group CBT interventions, in which effect size was moderate and slightly inferior to non-group intervention, which had a large effect size. Older individuals suffering from PD with psychological disorders are more likely to experience inferiority, stigma and fear because of increased dysfunction [29]. Patients in one group of older adults with PD tended to have unequal degrees of disease, and by mutually comparing themselves may have deepened their own withdrawal and enhanced communication difficulties. Moreover, conducting CBT in a group is not targeted enough compared with individual therapy where individuals may fully focus without the distraction of other participants, which makes the efficacy of group intervention limited. In our study, although the improvement in symptoms seen after group CBT intervention was indelible, the non-group CBT intervention had the most significant effect.

We also analyzed outcomes in different regions based on the nationality of the patients included in the studies. These patients were recruited in Europe, North America and Oceania. The effect sizes observed in both depression and anxiety were moderate to large, with no obvious differences among patients from these three continents. More specific research is needed to validate these results in populations across other geographic locations.

Sleep disorders are a high-morbidity-rate complication in PD that have a negative impact on patients [30]. Our results showed that CBT had a moderate overall effect on sleep disorders (*E* = -0.44). Our subgroup analysis showed no significant difference in the efficacy of different methods of CBT, including online or offline and group or non-group interventions, on sleep disorders. Because all forms of CBT intervention showed at least a moderate effect size, the intervention forms had no interactions with the results. We separately analyzed the change in insomnia post-intervention, which resulted in a large effect size (*E* = -0.94). It has reported CBT is beneficial to insomnia in patients with or without PD [31]. This may be because insomnia has been associated more with persistent psychological factors than specific disease characteristics in the general population [32], while the goal of CBT in treating insomnia is to alter erroneous perceptions, cognitive arousal and maladaptive behaviors toward sleep hygiene [33]. Another result revealed that sleep quality did not ameliorate, which demonstrates that CBT can influence sleep duration rather than sleep architecture. Only one set of data on daytime sleep was provided in one of the included studies, precluding the ability to run a separate analysis. The data showed daytime sleep was reduced after CBT.

Assessments of both sleep and fatigue are often performed simultaneously. We analyzed the effect of CBT on fatigue, which showed there was no evidence of a decrease in fatigue after CBT. Physical fatigue caused by dyskinesia that cannot be alleviated by CBT may be one of the reasons for the small improvement, suggesting that motor and non-motor symptoms potentially synergize, and treatment of a single symptom may undermine optimal efficacy. Interestingly, as the actions of the sleep homeostat, short periods of nocturnal sleep and may cause daytime sleep or fatigue, implying a connection exists between sleep and fatigue as symptoms [34]. We can infer that improvements can be detected simultaneously in these symptoms after CBT.

Our analysis showed that CBT was moderately efficacious on stress. Stress may aggravate symptoms of PD [35]. It has been reported that CBT relieves stress by promoting self-efficacy [36]. In addition, one article dealt with apathy and psychiatry rating and one with negative thoughts, and each of the three indicators had only a set of data. Limited data have shown that the intervention group had a more desired effect and that CBT improved outcomes compared to the control group.

Our results showed quality of life did not improve significantly for patients with PD after CBT. One of the articles had increased heterogeneity that may have been related to the inclusion of light therapy. Patients were allowed to read about the mechanisms of light therapy independently, and therefore they were not completely blinded to the intervention in this study. Several scales were used to assess the quality of life. The PDQ is a questionnaire with 39 questions in eight dimensions [37]. The PDQ contains many questions about physical problems and/or mobility difficulties that cannot be solved through psychological therapy, a possible reason for the lack of improvement in quality of life after CBT. We performed a subgroup analysis of the scales used to measure quality of life and found a moderate effect on quality of life in studies that did not use the PDQ, while there was no significant efficacy of CBT on quality of life in studies that used the PDQ. This may suggest that the choice of scales had an influence on the evaluation results of quality of life. We also found the duration of the intervention affected results, as group CBT interventions for > 10 weeks yielded moderate effect sizes on quality of life. Previous literature has reported that depression, anxiety, sleep disorder and apathy are predictors of quality of life [38]. However, in our study, although we observed an improvement in depression, anxiety and sleep disorder after CBT, quality of life did not particularly improve. The relationship between non-motor symptoms and quality of life in PD remains to be explored.

All of the studies in this analysis were RCTs, and appropriate processing and analyses were made for the missing data. The studies reported no serious adverse events. Our quantitative analysis revealed little heterogeneity among studies and after exploring possible reasons, the two papers causing the increased heterogeneity were removed from analysis. The scales used to measure the outcomes of the included studies are accredited and have been used extensively in clinical practice, such as the Hamilton Rating Scale (HAM), Beck Anxiety Inventory (BAI) and others. We chose the random effects model to eliminate the discrepancy between scales to ensure reliable results.

This analysis had several limitations. First, the sample sizes of the included studies were generally small, with the largest sample having only 80 participants, which may lead to a lack of generalizable results. We need wider clinical trials to perform further research. Second, inadequate data on apathy, negative thoughts and psychiatric rating prevented us from making a comprehensive assessment of these indexes. More relevant studies need to be included to obtain more reliable quantitative results. It is necessary to exploit more convenient and suitable CBT intervention methods for patients with PD. The effect of long-term CBT intervention on non-motor symptoms and quality of life needs further study. Finally, the measurement scales used in the selected studies were validated but highly varied, resulting in the possibility that an outcome might be exaggerated or minimized during the process of data merging. Gathering symptom data using the same scales across studies would result in better compatibility and potentially more accurate results.

## Conclusion

Cognitive behavioral therapy had a significant effect on depression, anxiety, stress, sleep disorders and some other non-motor symptoms in patients with PD, while it had no effect on fatigue or quality of life. The longer the intervention, the greater the effect. Symptom improvements after individual and face-to-face CBT interventions are much more remarkable than after group interventions. The use of CBT should be considered as a standard clinical treatment for non-motor symptoms of PD, and more convenient and efficient forms should be explored.

## Data Availability

We have confirmed.

## Acknowledgment

This work was funded by the National Natural Science Foundation (Nos. 81560059 and 81760058), Zhejiang Medical Health Science and Technology Project (No. 2019328893), the Scientific Research Fund of Shaoxing University (No. 20125025), and the National Training Program of Innovation and Entrepreneurship for College Students (No. 2017R10349001). We thank LetPub (www.letpub.com) for its linguistic assistance during the preparation of this manuscript.

---

**Fig. S1. Forest plot of random effects model meta-analysis of the effect of CBT on stress**

Abbreviations: CBT = cognitive behavioral therapy.

**Fig. S2. Forest plot of random effects model meta-analysis of the effect of CBT on fatigue**

Abbreviations: CBT = cognitive behavioral therapy.

**Fig. S3. Forest plot of random effects model meta-analysis of the effect of CBT on apathy**

Abbreviations: CBT = cognitive behavioral therapy.

**Fig. S4. Forest plot of random effects model meta-analysis of the effect of CBT on negative thought**

Abbreviations: CBT = cognitive behavioral therapy.

**Fig. S5. Forest plot of random effects model meta-analysis of the effect of CBT on psychiatry rating**

Abbreviations: CBT = cognitive behavioral therapy.

**Fig. S6. Forest plot of random effects model meta-analysis of the effect of CBT on subgroups of QOL**

Abbreviations: CBT = cognitive behavioral therapy; QOL = quality of life; PDQ = Parkinson’s Disease Questionnaire; A = the effect of intervention time; B = the effect of varied scales.

